# Cardiorespiratory Limits Exist in HFpEF Independent of Age and BMI

**DOI:** 10.64898/2026.11.14.26368115

**Authors:** Serafino LaGalbo, Madhu Gowda, Timothy Hess, Parag Tipnis, Farhan Raza, Danish Saleh

## Abstract

**Background:** Heart-failure with preserved ejection fraction (HFpEF) is a clinical syndrome marked by exercise intolerance associated with impairments in cardiac and respiratory performance. HFpEF is associated with age and weight; however, it remains unclear whether cardiorespiratory limitations exist independent of these comorbidities.

**Hypothesis:** Independent of age and BMI, there are cardiorespiratory limitations inherent to the HFpEF syndrome that have yet to be fully elicited.

**Methods:** A patient cohort with a clinical diagnosis of HFpEF (HFpEF; N = 38) and counterparts without identifiable cardiopulmonary disease (control; N = 21) underwent phenotyping with echocardiography and cardiopulmonary exercise testing with invasive hemodynamic monitoring.

**Results:** Study cohorts were similar in age, BMI and height (p=NS for all metrics). Body-surface area (BSA) (2.1 m^2^ v 1.9 m^2^, p < 0.05), atrial fibrillation prevalence (50 v 14%, p < 0.05), and average E/e’ (11.5 v 9.5, p < 0.05) were elevated within the HFpEF cohort. Peak VO_2_ (14.1 v 19.1 mL/Kg/min, p < 0.001) was also diminished among those with HFpEF. Resting (17.1 v 11.4mmHg, p < 0.001) and exercise wedge pressures (29.3 v 16.7mmHg, p < 0.001) were elevated whereas stroke volume (93.3 v 110.4 mL, p < 0.05) and cardiac output (Direct Fick; 8.0 v 9.8, p < 0.05) during exercise were diminished in HFpEF patients. FVC, FVC%, FEV1 and FEV1% were also significantly diminished in the HFpEF cohort.

**Conclusions:** Cardiorespiratory limitations exist as a feature of the HFpEF syndrome, independent of age and BMI. BSA remained associated with HFpEF in keeping with adiposity as a feature of pathophysiology.

## Introduction

Heart-failure with preserved ejection fraction (HFpEF) is an age-associated clinical syndrome marked by dyspnea and exertional intolerance. Clinically, patients with HFpEF are often sedentary, older, overweight or obese, and living with cardiovascular comorbidities including hypertension, diabetes, atrial fibrillation, and obstructive sleep apnea.^1,2^ The HFpEF syndrome may be invoked by pairing clinical symptoms with signs of heart failure that cannot be attributed to underlying valvular heart disease and significant systolic ventricular dysfunction. The diagnosis is further supported by echocardiographic or invasive catheter-based hemodynamic evidence of impairment in ventricular relaxation and elevated left-heart filling pressures at rest or with exercise.^3^ Biochemical evidence of myocardial muscle stretch and/or myocardial injury is not uncommon in the HFpEF syndrome.^4,5^

Impairments in respiratory performance parameters have also been observed as a feature of the HFpEF syndrome.^6–11^ Specifically, abnormal pulmonary function testing (PFTs), including diminished diffusion-capacity of the lungs for carbon monoxide (DLCO), and decrements in respiratory mechanics have been described in patient cohorts with HFpEF. Moreover, increased dead-space ventilation as a function chronic obstructive pulmonary disease (COPD) is a common HFpEF linked comorbidity.^12^

It remains to be understood whether the constellation of cardiorespiratory deficits in the HFpEF syndrome reflects intrinsic cardiopulmonary dysfunction or an age and body-size related phenomena. Seminal studies characterizing the HFpEF syndrome described clinical features without adjusting for age and body mass.^1,13^ Herein, we examine cardiorespiratory parameters in an age and body-mass index (BMI) restricted patient cohort to uncouple the aforenoted demographic elements from cardiorespiratory performance parameters.

## Methods

This retrospective cohort study sampled patients enrolled in the Pulmonary Hypertension Study Cohort (PHSC) at the University of Wisconsin – Madison. Enrollment included patients with a prior clinical diagnosis of HFpEF and counterparts without identifiable cardiopulmonary disease. The PHSC included patients with NYHA Class II-III exertional limitations and who were previously evaluated in the pulmonary hypertension clinic at the University of Wisconsin – Madison. Patients enrolled in the PHSC underwent phenotyping with echocardiography, pulmonary function testing, and rest-to-exercise cardiopulmonary exercise testing using a semi-recumbent ergometer and simultaneous right heart catheterization with expired gas analysis. Patients were designated as having WHO group 2 pulmonary hypertension or non-cardiac dyspnea based on hemodynamic evidence following the 2022 ESC/ERS Guidelines.^14^ Patients with LVEF <50%, supplemental oxygen use, serum creatinine >2 mg/dL or on dialysis, primary lung diseases with forced vital capacity <60% predicted, and severe valvular heart disease were excluded from the PHSC. For this study, patients over the age of 75 and those with a BMI > 35 were excluded to limit the impact of extreme phenotypes on study observations **(Figure 1).** The final study cohort included 38 individuals with HFpEF and pulmonary hypertension and 21 counterparts without identifiable cardiopulmonary disease (control; N = 21). The study was approved by the University of Wisconsin-Madison’s institutional review board.

**Figure 1:**
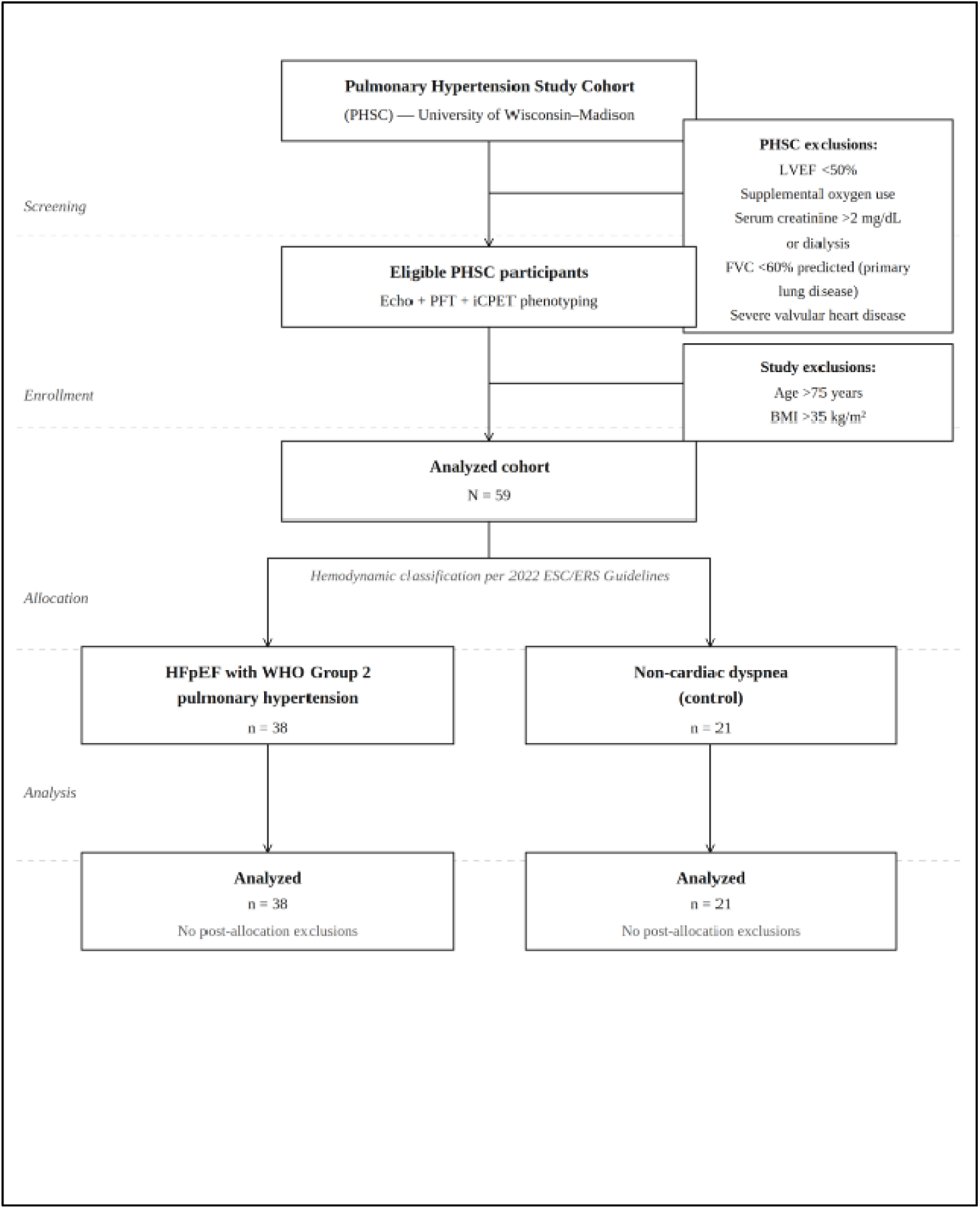
CONSORT diagram

### Data Analysis

Continuous measurements were summarized as means ± standard deviations while categorical measurements tabulated as counts and percentages. Comparisons between HFpEF and control groups were evaluated by Wilcoxon rank-sum tests for continuous measures and Chi-square or Fisher’s exact test for categorical values. A p-value of less than 0.05 was considered statistically significant. All analyses were conducted using SAS version 9.4.

## Results

There was no difference between the HFpEF and control arms with respect to average age, BMI, height, female sex-ratio, and history of tobacco use **(Table 1).** However, patients in the HFpEF arm had a higher body-surface area (BSA) when compared to their control counterparts (2.1 m^2^ 1.9 m^2^, p < 0.05).

**Table 1.**

| **Characteristic** | **Control (N = 21)** | **HFpEF (N = 38)** | **P Value** |
|---|---|---|---|
| Age, years | 64.5 ± 7.2 | 65.7 ± 8.6 | 0.345 |
| Female sex, no. (%) | 12 (57.1) | 19 (50.0) | 0.800 |
| BMI, kg/m² | 30.9 ± 4.2 | 30.9 ± 4.7 | 0.526 |
| Height, cm | 167.2 ± 11.7 | 173.2 ± 11.1 | 0.054 |
| BSA, m² | 1.9 ± 0.2 | 2.1 ± 0.3 | 0.017 |
| Current or former smoker, no. (%) | 8 (38.1) | 18 (47.4) | 0.680 |
| Hypertension, no. (%) | 12 (57.1) | 32 (84.2) | 0.048 |
| Hyperlipidemia, no. (%) | 16 (76.2) | 24 (64.9) | 0.548 |
| Atrial fibrillation, no. (%) | 3 (14.3) | 19 (50.0) | 0.015 |
| Diabetes mellitus, no. (%) | 4 (19.0) | 12 (31.6) | 0.465 |
| Chronic kidney disease, no. (%) | 3 (14.3) | 13 (35.1) | 0.161 |
| Creatinine, mg/dL | 1.1 ± 0.5 | 1.4 ± 0.7 | 0.060 |
| Obstructive sleep apnea, no. (%) | 12 (57.1) | 20 (54.1) | 1.000 |
| Prior cardiac surgery, no. (%) | 2 (9.5) | 7 (18.4) | 0.595 |
| AV nodal blocker, no. (%) | 8 (38.1) | 25 (65.8) | 0.075 |
| ACE inhibitor/ARB/ARNi, no. (%) | 5 (23.8) | 15 (39.5) | 0.352 |
| Mineralocorticoid receptor antagonist, no. (%) | 5 (23.8) | 11 (28.9) | 0.905 |
| SGLT2 inhibitor, no. (%) | 1 (4.8) | 13 (34.2) | 0.026 |
| Diuretic, no. (%) | 6 (28.6) | 26 (68.4) | 0.008 |

*Data presented as mean ± SD or number (%). ACE = angiotensin-converting enzyme; ARB = angiotensin II receptor blocker; ARNi = angiotensin receptor–neprilysin inhibitor; AV = atrioventricular; BMI = body mass index; BSA = body surface area; HFpEF = heart failure with preserved ejection fraction; SGLT2i = sodium-glucose cotransporter-2 inhibitor.*

Patients in the HFpEF study arm were more likely to have hypertension (84.2% v 57.1%, p < 0.05) and atrial fibrillation (50.0% v 14.3%, p < 0.05). Diabetes (31.6% v 19.0%, p 0.465) and higher-baseline creatinine (1.4 v 1.1, p 0.060) were more prevalent in the HFpEF cohort; however, these differences were not statistically significant. There was no difference between the two arms with respect to hyperlipidemia and obstructive sleep apnea.

Patients in the HFpEF arm were more likely to be managed with a diuretic (68.4 v 28.6, p < 0.05) or a sodium-glucose transporter 2 inhibitor (SGLT2i) (34.2% v 4.8%, p < 0.05). Atrio-ventricular (AV) nodal blocker therapy was observed more often among patients with HFpEF (65.8% v 38.1%, p 0.075); however, this did not meet criteria for statistical significance. There was no difference between the study groups with respect to angiotensin-converting enzyme (ACE)/angiotensin-receptor blocker (ARB)/Angiotensin-receptor blocker with neprilysin inhibitor (ARNi) compound use, mineralocorticoid receptor antagonist (MRA) use, and/or prior cardiac surgery.

The forced vital capacity (FVC) (2.8 L v 3.9 L, p < 0.05), FVC% (75.8 v 100.6, p < 0.05), forced expiratory volume (FEV) (1.9 L v 2.8 L, p < 0.05), and FEV% (68.5 v 98.4, p < 0.05) were diminished in the HFpEF arm compared to the control arm **(Table 2**, **Figure 2)**. The ratio of FEV1/FVC was not different between the study arms. Residual lung volume (RV) (2.2 L v 1.9 L, p 0.060), RV% (102.5 v 98.7, p = 0.185), lung diffusion capacity (DLCO%) (68 v 87.2, p = 0.063) and lung diffusion capacity as a ratio of alveolar volume (DLCO/VA) (89.3 v 76.2, p 0.168) were lower in patients with HFpEF but these factors did not meet threshold for statistical significance. There was no difference in total lung capacity (TLC) between the two groups; however, TLC% trended down in patients with HFpEF (90.7 v 100.9, p < 0.123).

**Figure 2:**
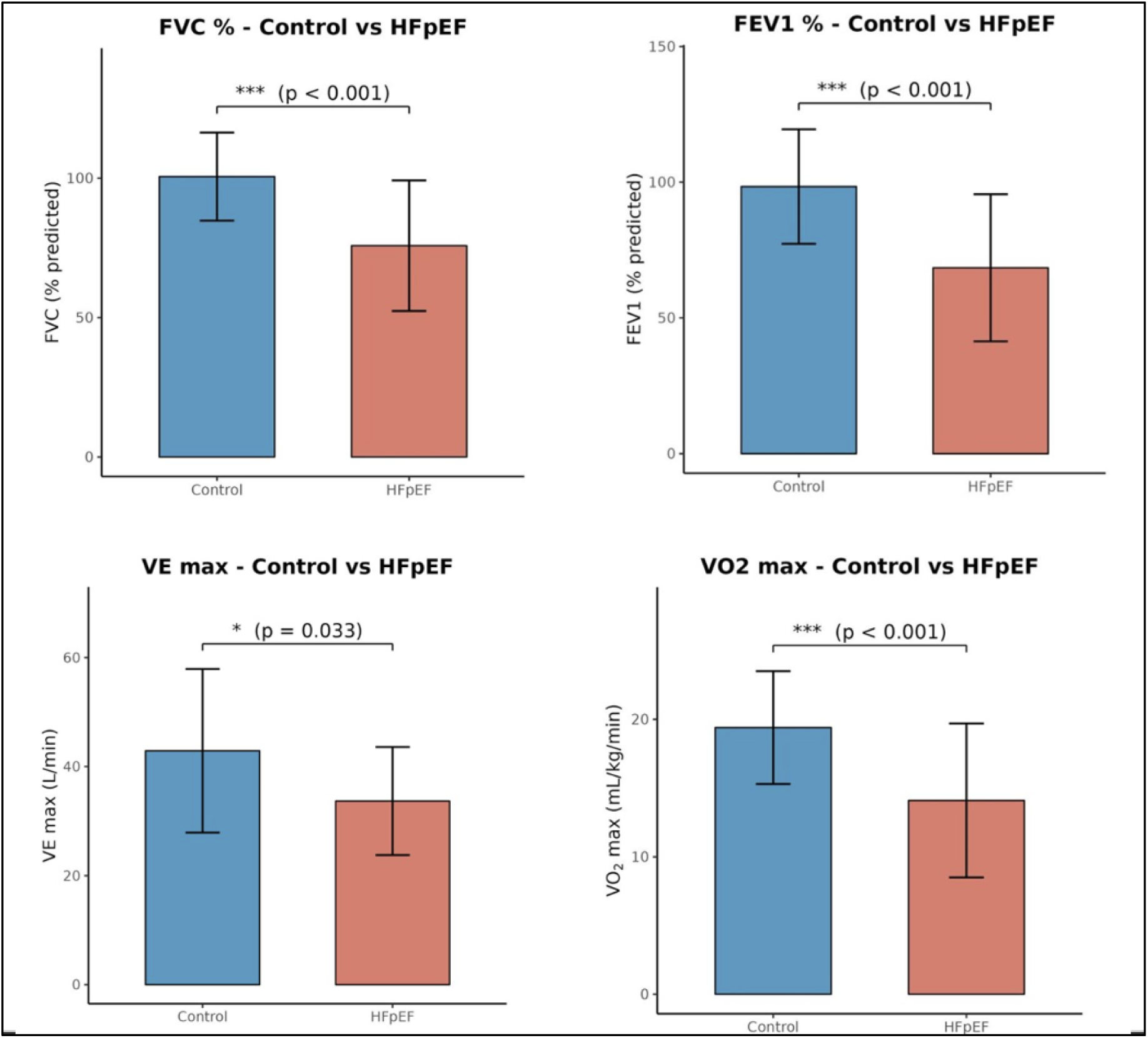
Comparison of percent predicted forced vital capacity (FVC%), expiratory volume in 1 second % predicted (FEV1 %), maximal oxygen consumption (VO_2_ max) and maximal ventilation (VE max) between HFpEF and control groups.

**Table 2:** Pulmonary Function Data.

| **Characteristic** | **Control (N = 21)** | **HFpEF (N = 38)** | **P Value** |
|---|---|---|---|
| FVC, L | 3.7 ± 0.9 | 2.8 ± 1.0 | 0.002 |
| FVC, % predicted | 100.6 ± 15.8 | 75.8 ± 23.4 | <0.001 |
| FEV1, L | 2.8 ± 0.7 | 1.9 ± 0.9 | 0.001 |
| FEV1, % predicted | 98.4 ± 21.1 | 68.5 ± 27.1 | <0.001 |
| FEV1/FVC | 74.2 ± 8.4 | 62.0 ± 13.1 | <0.001 |
| TLC, L | 5.4 ± 1.1 | 5.0 ± 1.4 | 0.322 |
| TLC, % predicted | 100.9 ± 19.8 | 90.7 ± 24.3 | 0.123 |
| RV, L | 1.9 ± 0.4 | 2.2 ± 0.7 | 0.060 |
| RV, % predicted | 98.7 ± 16.5 | 102.5 ± 32.0 | 0.185 |
| DLCO, % predicted | 87.2 ± 20.5 | 68.0 ± 29.5 | 0.003 |
| DLCO/VA, % predicted | 89.3 ± 30.8 | 76.2 ± 34.9 | 0.168 |

*Data presented as mean ± SD. DLCO = diffusing capacity of the lungs for carbon monoxide; FEV1 = forced expiratory volume in 1 second; FVC = forced vital capacity; HFpEF = heart failure with preserved ejection fraction; RV = residual volume; TLC = total lung capacity; VA = alveolar volume.*

Left-ventricular ejection fraction (LVEF), left-ventricular end-diastolic diameter (LVEDd), and the left-ventricular velocity-time-integral (LVOT VTI) were similar when comparing patients with HFpEF to control counterparts **(Table 3).** The mitral inflow velocity (E) (88.2 v 70.8 cm/s, p < 0.05), average mitral inflow velocity divided by mitral annular tissue velocity (E/e’) (11.5 v 9.5, p < 0.05), left-atrial volume indexed (LAVI) (39.9 v 30.0 mL/m^2, p < 0.05), pulmonary artery systolic pressure (PASP) (44.5 mmHg v 30.4 mmHg, p < 0.05), and grade of tricuspid regurgitation (TR) (1.5 v 0.8, p < 0.05) were greater in patients with HFpEF.

**Table 3:**
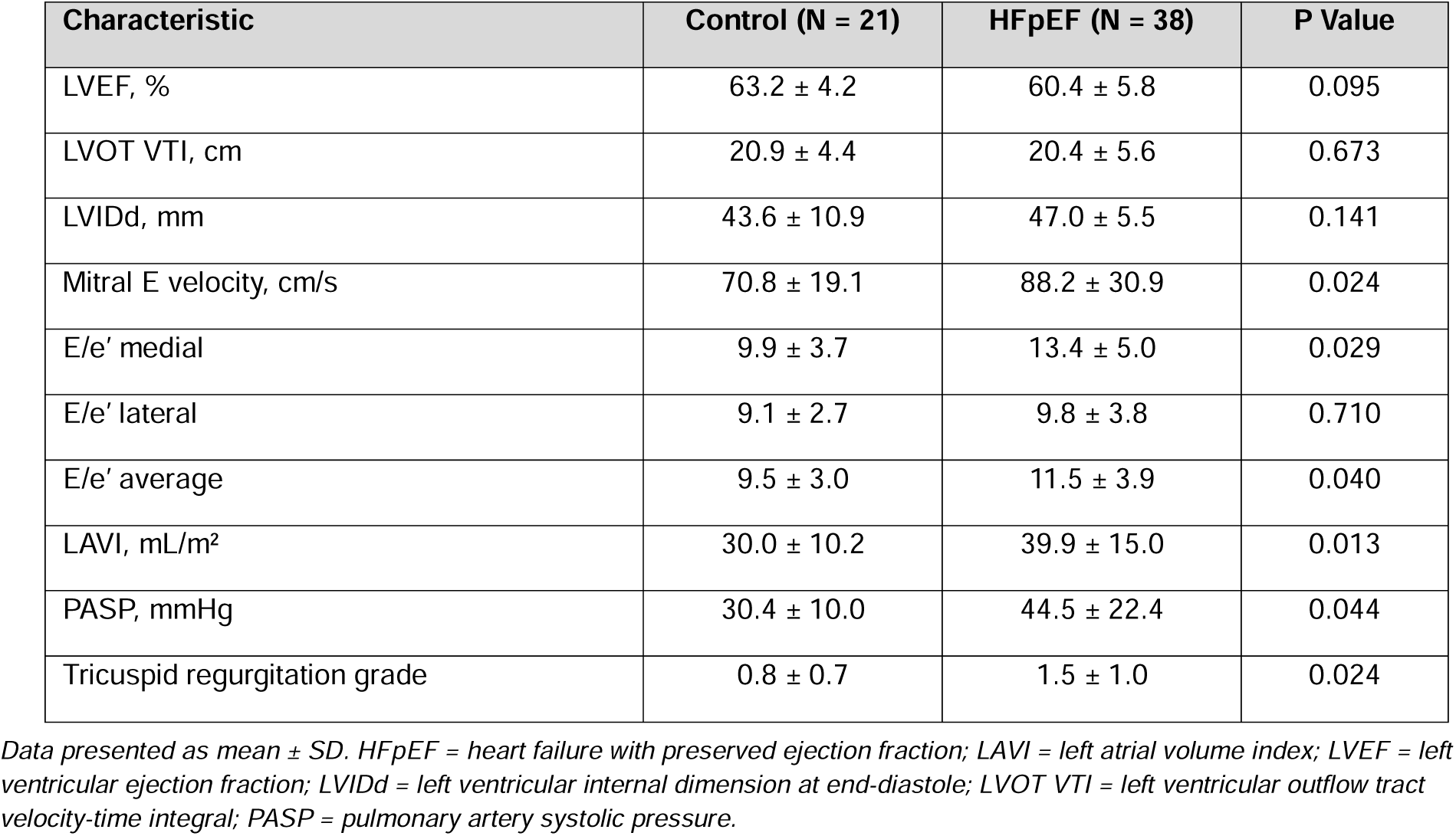
Transthoracic Echocardiography Data.

| **Characteristic** | **Control (N = 21)** | **HFpEF (N = 38)** | **P Value** |
|---|---|---|---|
| LVEF, % | 63.2 ± 4.2 | 60.4 ± 5.8 | 0.095 |
| LVOT VTI, cm | 20.9 ± 4.4 | 20.4 ± 5.6 | 0.673 |
| LVIDd, mm | 43.6 ± 10.9 | 47.0 ± 5.5 | 0.141 |
| Mitral E velocity, cm/s | 70.8 ± 19.1 | 88.2 ± 30.9 | 0.024 |
| E/e’ medial | 9.9 ± 3.7 | 13.4 ± 5.0 | 0.029 |
| E/e’ lateral | 9.1 ± 2.7 | 9.8 ± 3.8 | 0.710 |
| E/e’ average | 9.5 ± 3.0 | 11.5 ± 3.9 | 0.040 |
| LAVI, mL/m<sup>2</sup> | 30.0 ± 10.2 | 39.9 ± 15.0 | 0.013 |
| PASP, mmHg | 30.4 ± 10.0 | 44.5 ± 22.4 | 0.044 |
| Tricuspid regurgitation grade | 0.8 ± 0.7 | 1.5 ± 1.0 | 0.024 |

*Data presented as mean ± SD. HFpEF = heart failure with preserved ejection fraction; LAVI = left atrial volume index; LVEF = left ventricular ejection fraction; LVIDd = left ventricular internal dimension at end-diastole; LVOT VTI = left ventricular outflow tract velocity-time integral; PASP = pulmonary artery systolic pressure.*

Right-sided cardiac pressures were elevated at rest and with exercise in patients with HFpEF compared to control counterparts ((RV systolic pressure (RVSP), rest, 47.2 mmHg v 31.3 mmHg, p < 0.001; RVSP, exercise, 74.8 mmHg v 42.5 mmHg, p < 0.001; pulmonary artery systolic pressure (sPAP), rest, 47.7 mmHg v 29.6 mmHg, p < 0.001; sPAP, exercise, 78.3 mmHg v 46.4 mmHg, p < 0.001)) (**Table 4)**.

**Table 4:**
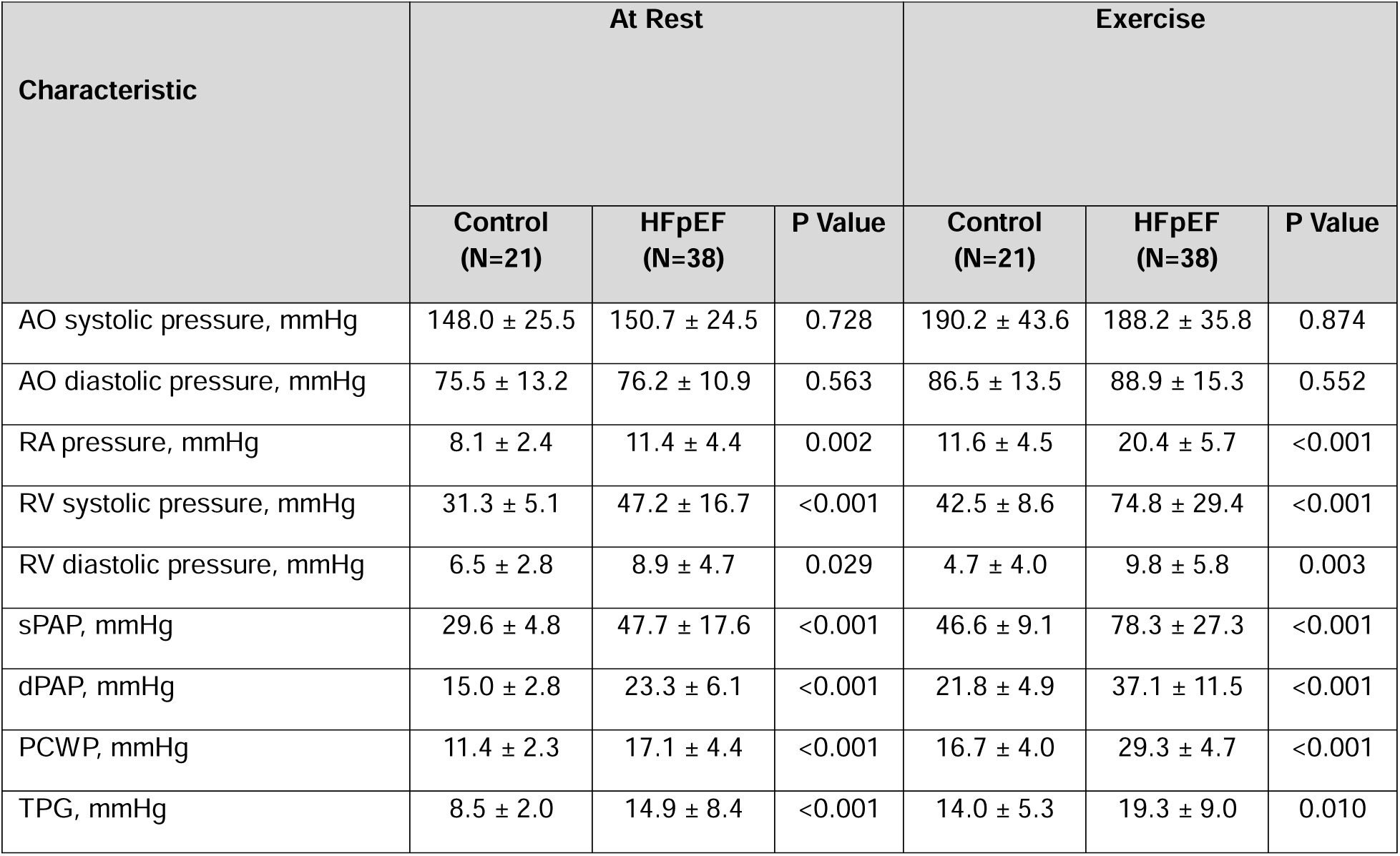
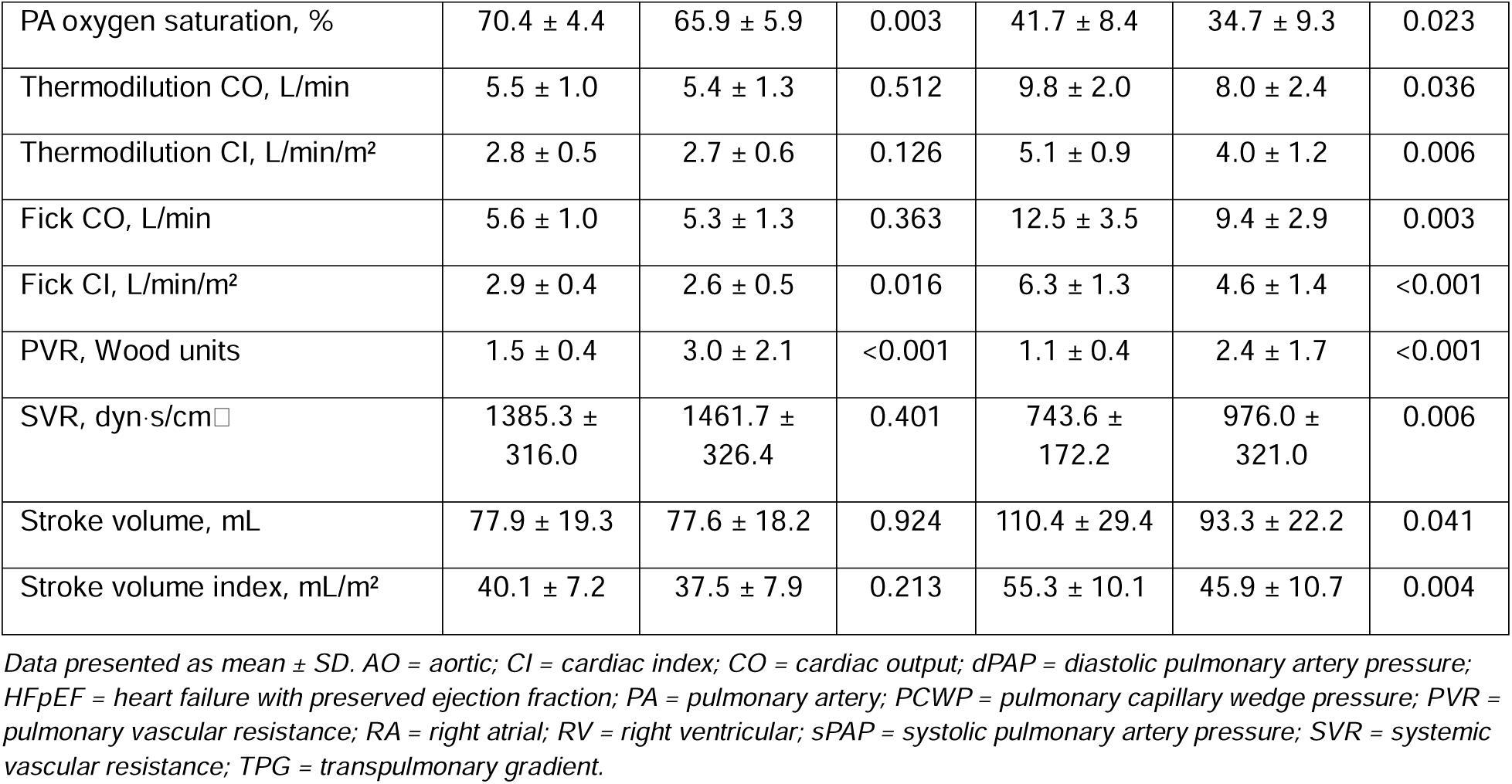
Invasive Hemodynamic Data at Rest and During Exercise.

| **Characteristic** | **At Rest** |  |  | **Exercise** |  |  |
|---|---|---|---|---|---|---|
|  | **Control (N=21)** | **HFpEF (N=38)** | **P Value** | **Control (N=21)** | **HFpEF (N=38)** | **P Value** |
| AO systolic pressure, mmHg | 148.0 ± 25.5 | 150.7 ± 24.5 | 0.728 | 190.2 ± 43.6 | 188.2 ± 35.8 | 0.874 |
| AO diastolic pressure, mmHg | 75.5 ± 13.2 | 76.2 ± 10.9 | 0.563 | 86.5 ± 13.5 | 88.9 ± 15.3 | 0.552 |
| RA pressure, mmHg | 8.1 ± 2.4 | 11.4 ± 4.4 | 0.002 | 11.6 ± 4.5 | 20.4 ± 5.7 | <0.001 |
| RV systolic pressure, mmHg | 31.3 ± 5.1 | 47.2 ± 16.7 | <0.001 | 42.5 ± 8.6 | 74.8 ± 29.4 | <0.001 |
| RV diastolic pressure, mmHg | 6.5 ± 2.8 | 8.9 ± 4.7 | 0.029 | 4.7 ± 4.0 | 9.8 ± 5.8 | 0.003 |
| sPAP, mmHg | 29.6 ± 4.8 | 47.7 ± 17.6 | <0.001 | 46.6 ± 9.1 | 78.3 ± 27.3 | <0.001 |
| dPAP, mmHg | 15.0 ± 2.8 | 23.3 ± 6.1 | <0.001 | 21.8 ± 4.9 | 37.1 ± 11.5 | <0.001 |
| PCWP, mmHg | 11.4 ± 2.3 | 17.1 ± 4.4 | <0.001 | 16.7 ± 4.0 | 29.3 ± 4.7 | <0.001 |
| TPG, mmHg | 8.5 ± 2.0 | 14.9 ± 8.4 | <0.001 | 14.0 ± 5.3 | 19.3 ± 9.0 | 0.010 |
| PA oxygen saturation, % | 70.4 ± 4.4 | 65.9 ± 5.9 | 0.003 | 41.7 ± 8.4 | 34.7 ± 9.3 | 0.023 |
| Thermodilution CO, L/min | 5.5 ± 1.0 | 5.4 ± 1.3 | 0.512 | 9.8 ± 2.0 | 8.0 ± 2.4 | 0.036 |
| Thermodilution CI, L/min/m<sup>2</sup> | 2.8 ± 0.5 | 2.7 ± 0.6 | 0.126 | 5.1 ± 0.9 | 4.0 ± 1.2 | 0.006 |
| Fick CO, L/min | 5.6 ± 1.0 | 5.3 ± 1.3 | 0.363 | 12.5 ± 3.5 | 9.4 ± 2.9 | 0.003 |
| Fick CI, L/min/m<sup>2</sup> | 2.9 ± 0.4 | 2.6 ± 0.5 | 0.016 | 6.3 ± 1.3 | 4.6 ± 1.4 | <0.001 |
| PVR, Wood units | 1.5 ± 0.4 | 3.0 ± 2.1 | <0.001 | 1.1 ± 0.4 | 2.4 ± 1.7 | <0.001 |
| SVR, dyn·s/cm□ | 1385.3 ± 316.0 | 1461.7 ± 326.4 | 0.401 | 743.6 ± 172.2 | 976.0 ± 321.0 | 0.006 |
| Stroke volume, mL | 77.9 ± 19.3 | 77.6 ± 18.2 | 0.924 | 110.4 ± 29.4 | 93.3 ± 22.2 | 0.041 |
| Stroke volume index, mL/m<sup>2</sup> | 40.1 ± 7.2 | 37.5 ± 7.9 | 0.213 | 55.3 ± 10.1 | 45.9 ± 10.7 | 0.004 |

*Data presented as mean ± SD. AO = aortic; CI = cardiac index; CO = cardiac output; dPAP = diastolic pulmonary artery pressure; HFpEF = heart failure with preserved ejection fraction; PA = pulmonary artery; PCWP = pulmonary capillary wedge pressure; PVR = pulmonary vascular resistance; RA = right atrial; RV = right ventricular; sPAP = systolic pulmonary artery pressure; SVR = systemic vascular resistance; TPG = transpulmonary gradient.*

Left-sided cardiac pressures were elevated at rest and with exercise in the setting of HFpEF compared to the control arm (pulmonary-capillary wedge pressure (PCWP), rest 17.1 mmHg v 11.4 mmHg, p < 0.001; PCWP, exercise 29.3 mmHg v 16.7 mmHg, p < 0.001). The resting thermodilution cardiac index (CI TD) trended lower in patients with HFpEF (2.7 v 2.8, p = 0.12) whereas the resting cardiac index by direct Fick (CD DF) was reduced in patients with HFpEF (2.6 v 2.9, p < 0.05). During exercise, each of CI TD (4.0 v 4.1, p < 0.05) and CI DF (4.6 v 6.3, p < 0.001) was reduced in HFpEF compared to control (**Figure 3).** In HFpEF patients, the cardiac output determined by thermodilution (CO TD) and direct Fick (CO DF) were not different in the resting condition but diminished during exercise (CO TD, exercise, 8.0 L/min v 9.8 L/min, p < 0.05; CO DF, exercise, 12.5 L/min v 9.4 L/min, p < 0.05) (**Figure 4).**

**Figure 3:**
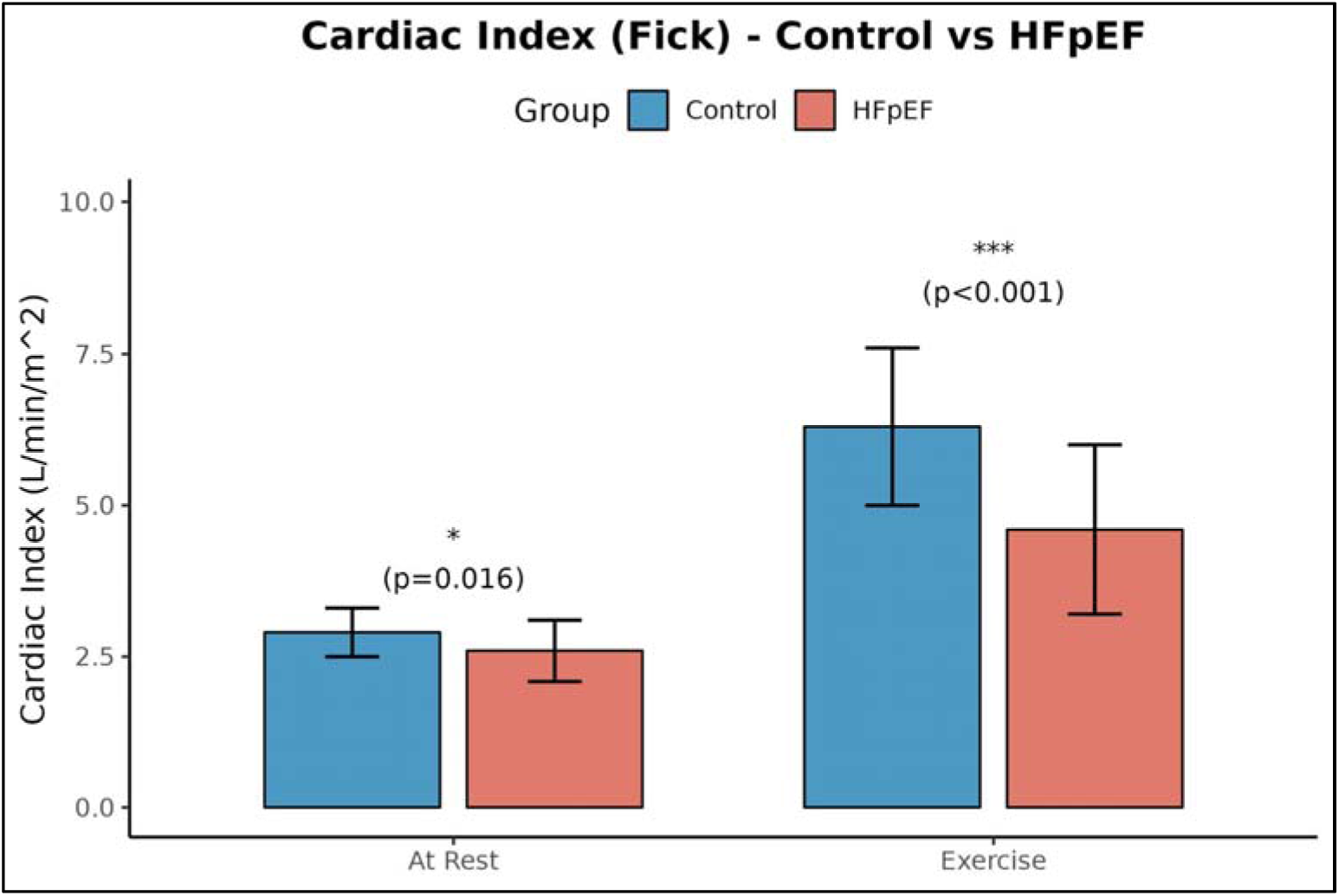
Comparison of cardiac index (Fick) at rest and exercise between HFpEF and control groups.

**Figure 4:**
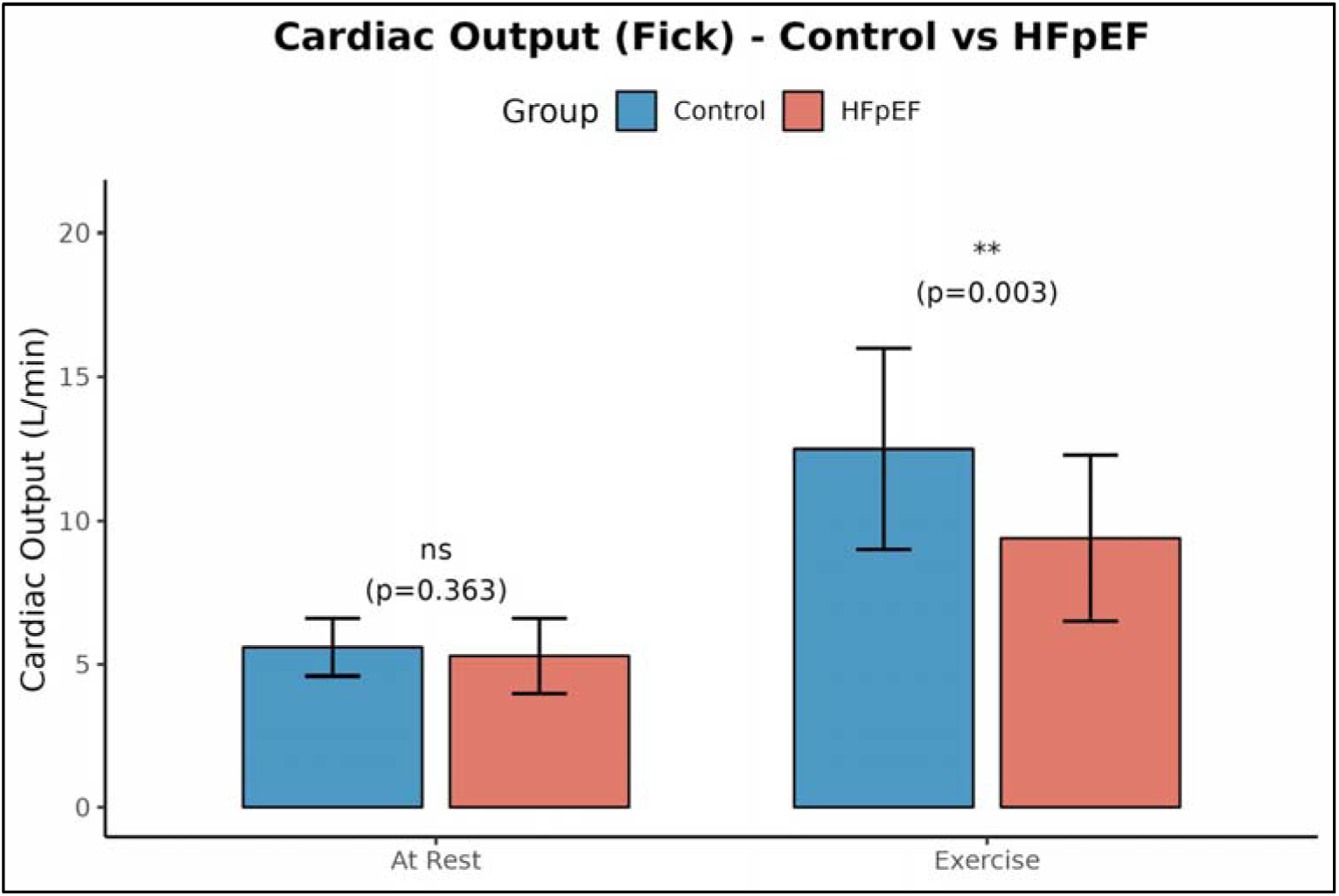
Comparison of cardiac output (Fick) at rest and exercise between HFpEF and control groups.

Differences in the resting and exercise associated stroke-volume and indexed stroke volume were analogous to changes in the cardiac output and index as determined by thermodilution. The resting and exercise-associated pulmonary-artery saturation (rest, 65.9 v 70.4, p < 0.05; exercise, 34.7 v 41.7, p < 0.05) were reduced in HFpEF compared to the control condition. The transpulmonary gradient (TPG) (rest, 14.9 v 8.5, p < 0.001; exercise, 19.3 v 14.0, p < 0.05) was increased in patients with HFpEF whereas systemic vascular resistance (SVR) was only increased during exercise (976 dyn•s/cm^5^ v 743 dyn•s/cm^5^, p < 0.05) in patients with HFpEF. Resting and exercise-associated aortic pressures were similar in both study arms (**Table 4)**.

In exercise testing, patients in the HFpEF cohort performed less work (60.8 w v 89.8 w, p < 0.05) and although patients with HFpEF trended towards less exercise time (7.2 min v 8.7 min, p = 0.21), this difference was not significant **(Table 5)**. The peak exercise-associated heart rate was blunted in patients with HFpEF (98.2 bpm v 114.4 bpm, p < 0.05) (**Figure 5)**. Each of maximal exercise ventilation (VE max) and peak VO_2_ (VO_2_ max) were reduced in the HFpEF cohort (VE Max: 33.7 L/min v 42.9 L/min, p < 0.05; VO_2_ max: 14.1ml/kg/min v 19.4 ml/kg/min, p < 0.001) compared to controls (**Figure 2).** There was no difference between the groups with respect to the respiratory exchange ratio (RER), minute ventilation to CO_2_ production (VE/VCO_2_) ratios at rest during exercise, the VE/CO_2_ slope, end-tidal CO_2_ at rest and during exercise, or the oxygen uptake efficiency slope (OUES).

**Figure 5:**
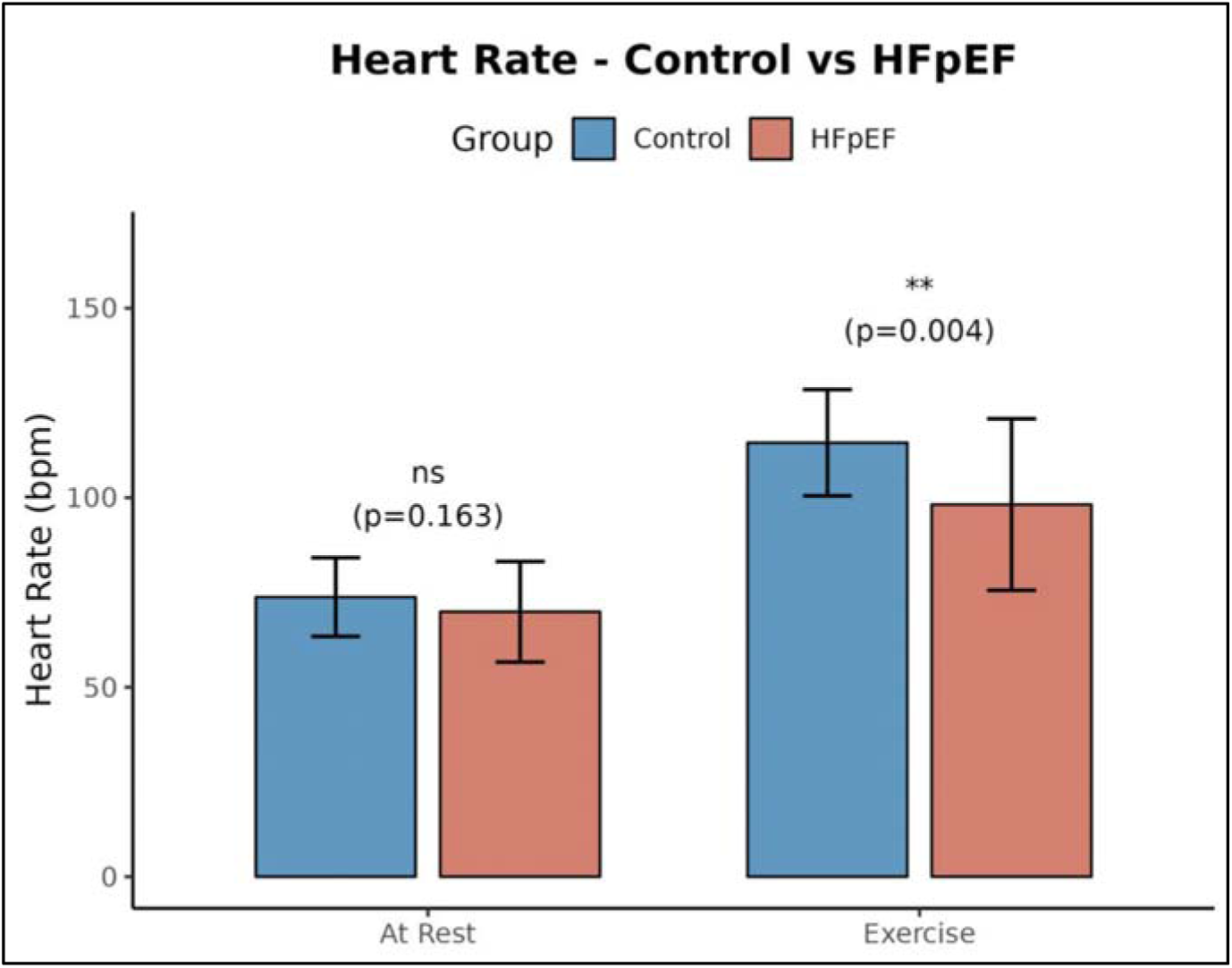
Comparison of heart rate at rest and exercise between HFpEF and control groups.

**Table 5a/b.**
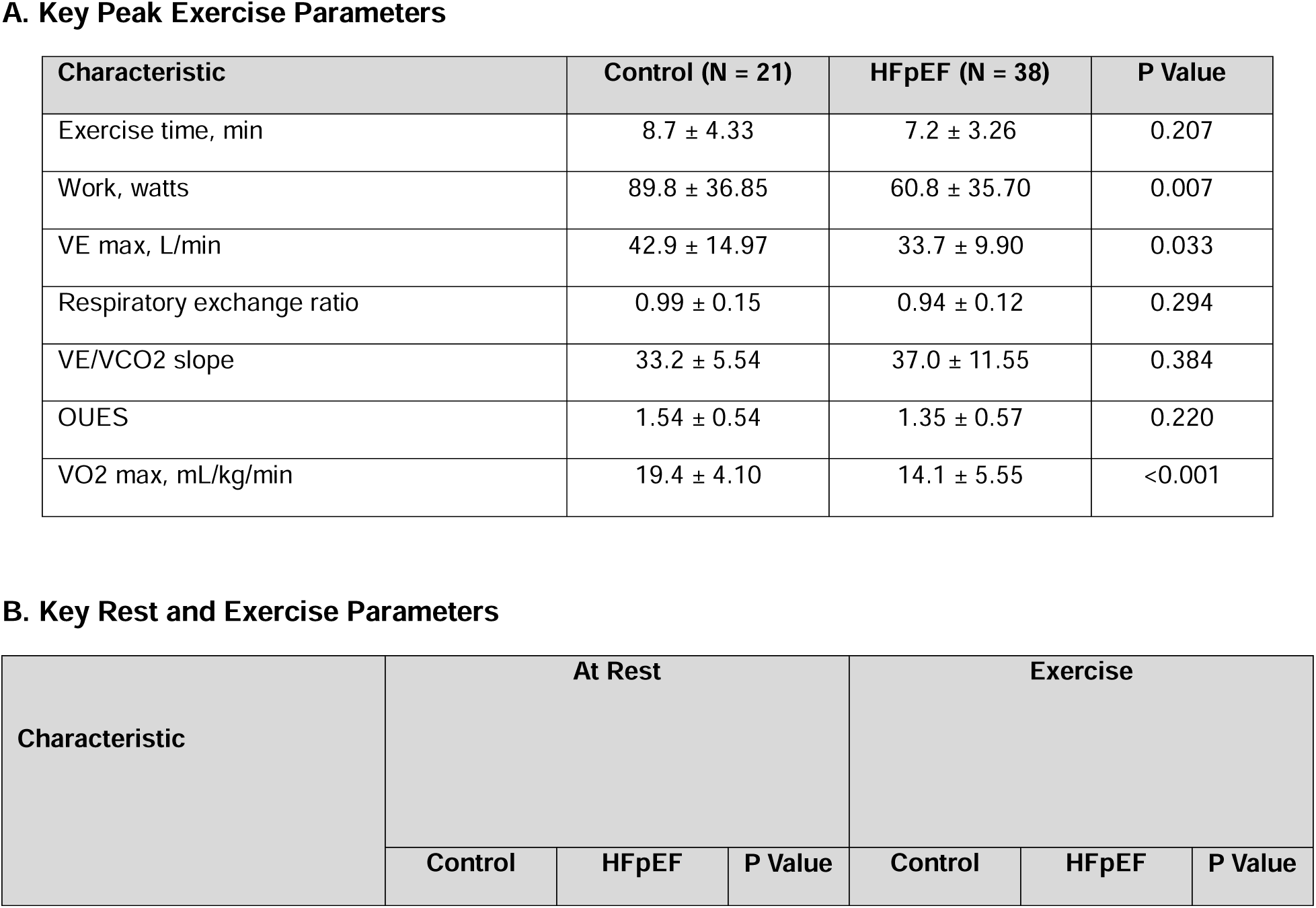
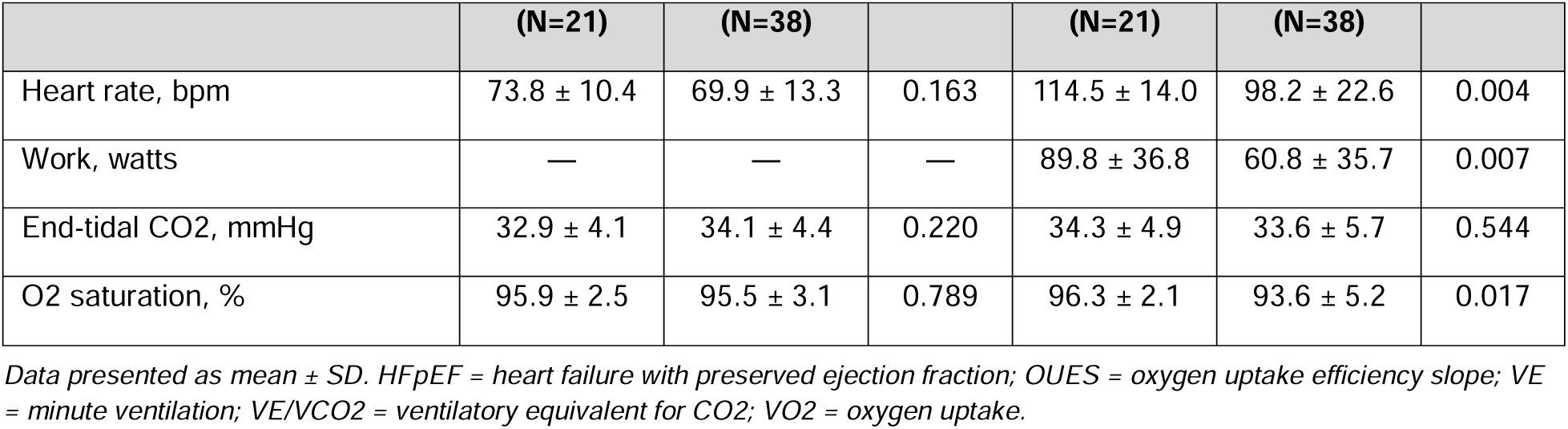

| A. Key Peak Exercise Parameters |  |  |  |  |  |  |
| Characteristic | Control (N = 21) | HFpEF (N = 38) | P Value |  |  |  |
|---|---|---|---|---|---|---|
| Exercise time, min | 8.7 ± 4.33 | 7.2 ± 3.26 | 0.207 |  |  |  |
| Work, watts | 89.8 ± 36.85 | 60.8 ± 35.70 | 0.007 |  |  |  |
| VE max, L/min | 42.9 ± 14.97 | 33.7 ± 9.90 | 0.033 |  |  |  |
| Respiratory exchange ratio | 0.99 ± 0.15 | 0.94 ± 0.12 | 0.294 |  |  |  |
| VE/VCO2 slope | 33.2 ± 5.54 | 37.0 ± 11.55 | 0.384 |  |  |  |
| OUES | 1.54 ± 0.54 | 1.35 ± 0.57 | 0.220 |  |  |  |
| VO2 max, mL/kg/min | 19.4 ± 4.10 | 14.1 ± 5.55 | <0.001 |  |  |  |
| **B. Key Rest and Exercise Parameters** |  |  |  |  |  |  |
| **Characteristic** | **At Rest** |  |  | **Exercise** |  |  |
|  | **Control** | **HFpEF** | **P Value** | **Control** | **HFpEF** | **P Value** |
|  | **(N=21)** | **(N=38)** |  | **(N=21)** | **(N=38)** |  |
| Heart rate, bpm | 73.8 ± 10.4 | 69.9 ± 13.3 | 0.163 | 114.5 ± 14.0 | 98.2 ± 22.6 | 0.004 |
| Work, watts | — | — | — | 89.8 ± 36.8 | 60.8 ± 35.7 | 0.007 |
| End-tidal CO2, mmHg | 32.9 ± 4.1 | 34.1 ± 4.4 | 0.220 | 34.3 ± 4.9 | 33.6 ± 5.7 | 0.544 |
| O2 saturation, % | 95.9 ± 2.5 | 95.5 ± 3.1 | 0.789 | 96.3 ± 2.1 | 93.6 ± 5.2 | 0.017 |

*Data presented as mean ± SD. HFpEF = heart failure with preserved ejection fraction; OUES = oxygen uptake efficiency slope; VE = minute ventilation; VE/VCO2 = ventilatory equivalent for CO2; VO2 = oxygen uptake.*

## Discussion

Herein, we examine the cardiopulmonary features of an age and BMI restricted HFpEF cohort during rest and with exercise. We observe that independent of age and BMI, the HFpEF syndrome is associated with multiple cardiorespiratory limitations that may contribute to the experience of dyspnea and exercise intolerance. Of note, our results demonstrate that high left-sided filling and pulmonary capillary wedge pressures are not solely a limiting factor for exercise performance in these patients, which has also been demonstrated in previous studies.^15–17^ Specifically, our study finds that patients with HFpEF have impairments in pulmonary function parameters (FEV, FVC) and cardiac hemodynamics (heart rate, cardiac output) with elevations in right and left-sided pressures, all of which likely contribute to impairments in peak VO_2_ and ability to perform work.

This study reveals key clinical considerations for the evaluation of dyspnea and exertional intolerance pertinent to the HFpEF syndrome. First off, the HFpEF syndrome is more commonly associated with atrial arrhythmias and, thus, patients with HFpEF are likely to be managed with AV nodal agents which likely dampens physiologic heart-rate responses associated with exertion.^18^ Conceivably, excess AV-nodal therapy may impair exercise capacity in an iatrogenic manner. Clinical decision-making to prioritize rhythm control strategies may be key to improving exercise tolerance in this patient population, as up-front rhythm control strategies have been associated with reduction in cardiovascular outcomes.^19^ Secondly, despite restricting our study cohort to a BMI < 35 with non-significant differences in BMI and height between the two groups, patients within the HFpEF arm had a larger BSA compared to their control counterparts. This suggests increased adiposity in the HFpEF group. This observation is consistent with the hypothesis that adiposity and its associated proinflammatory and profibrotic adipokines are a driver of the HFpEF syndrome that may serve as the seed of cardiopulmonary dysfunction even after excluding age and body mass.^20,21^ Not surprisingly, weight-loss and loss of adiposity have been associated with improved outcomes in this patient population.^22^

Finally, we observed reduced pulmonary artery oxygen saturation at rest and with exertion in the HFpEF cohort. This finding is suggestive of increased systemic oxygen metabolic need in the setting of increased adiposity. And, it is possible that patients with the HFpEF syndrome may experience impaired exercise tolerance as a function of an increased proportion of oxygen-consuming metabolic or adipose tissue.

Some limitations to our study should be acknowledged. Foremost, our study cohort is small, and patient data is limited to a single tertiary care academic center. Sample and patient selection biases inherent to small single center studies may limit the generalizability of our observations. Additionally, due to small sample size, the study was not sufficiently powered to perform broad statistical adjustments for possible confounding variables. For example, BSA remains a key difference between our HFpEF study cohort and control counterparts. This difference was tolerated for the study under the assumption that increased adiposity is a feature of the HFpEF syndrome as opposed to a modifying factor. Despite the aforenoted limitations, our study offers extensive phenotyping with observations that are consistent with the prior literature, altogether suggesting high fidelity.

## Conclusions

The experience of dyspnea and exercise intolerance of the HFpEF syndrome are likely multi-factorial in etiology. Patients with HFpEF tend to be overweight and/or obese and older and exertional intolerance can be a direct function of each of these factors. Of note, our study finds that the HFpEF syndrome features cardiorespiratory performance limitations that are likely independent of these demographic elements. Additional considerations in this patient population relate to increased oxygen-needs associated with increased adiposity and blunted exercise-associated heart-rate response in the setting of atrioventricular nodal blocker use. The examination of large highly phenotyped cohorts will be instrumental to understanding the origin(s) of dyspnea and exertional intolerance in HFpEF patients.

## Data Availability

All data produced in the present work are contained in the manuscript

## Notes

### Competing Interest Statement

The authors have declared no competing interest.

### Author Declarations

The IRB of the University of Wisconsin-Madison gave ethical approval for this work.

